# Differential immunogenicity of homologous versus heterologous boost in Ad26.COV2.S vaccine recipients

**DOI:** 10.1101/2021.10.14.21264981

**Authors:** Nicholas Khoo Kim Huat, Joey Ming Er Lim, Upkar S. Gill, Ruklanthi de Alwis, Nicole Tan, Justin Zhen Nan Toh, Jane E. Abbott, Carla Usai, Eng Eong Ooi, Jenny Guek Hong Low, Nina Le Bert, Patrick T. F. Kennedy, Antonio Bertoletti

## Abstract

Protection offered by COVID-19 vaccines wanes over time, requiring an evaluation of different boosting strategies to revert such a trend and enhance the quantity and quality of Spike-specific humoral and cellular immune responses. These immunological parameters in homologous or heterologous vaccination boosts have thus far been studied for mRNA and ChAdOx1 nCoV-19 vaccines, but knowledge on individuals who received a single dose of Ad26.COV2.S is lacking.

We studied Spike-specific humoral and cellular immunity in Ad26.COV2.S vaccinated individuals (n=55) who were either primed with Ad26.COV2.S only (n=13), or boosted with a homologous (Ad26.COV2.S, n=28) or heterologous (BNT162b2, n=14) second dose. We compared our findings with the results found in individuals vaccinated with a single (n=16) or double (n=44) dose of BNT162b2. We observed that a strategy of heterologous vaccination enhanced the quantity and breadth of both, Spike-specific humoral and cellular immunity in Ad26.COV2.S vaccinated. In contrast, the impact of homologous boost was quantitatively minimal in Ad26.COV2.S vaccinated and Spike-specific antibodies and T cells were narrowly focused to the S1 region. Although a direct association between quantity and quality of immunological parameters and in vivo protection has not been demonstrated, the immunological features of Spike-specific humoral and cellular immune responses support the utilization of a heterologous strategy of vaccine boost in individuals who received Ad26.COV2.S vaccination.

## Introduction

Vaccination has been the key strategy to reduce the incidence of SARS-CoV-2 infection and to protect from severe COVID-19 world-wide. Accelerated vaccine development efforts led to the approval of SARS-CoV-2 vaccines utilizing several different technological platforms, that displayed varying clinical efficacy with highest being associated with adenoviral vector and mRNA-based vaccines (1–5). Vaccine-induced protective efficacy is associated with their ability to induce neutralizing anti-Spike antibodies and Spike-specific T cells (3, 6, 7). Unfortunately, the appearance of the Delta variant and the progressive waning of antibody titers observed over time has reduced the protective efficacy of COVID-19 vaccines (5). These findings have ignited a debate about the need for possible booster vaccinations.

Ad26.COV2 (Johnson & Johnson) is a single dose vaccine (8) with protective efficacy against severe disease (4, 9). Single immunization with Ad26.COV2.S induced rapidly cellular immune responses as well as binding and neutralizing antibodies, including induction of RBD-specific binding antibodies in 90% of vaccine recipients (4, 9, 10). A recent report also indicated that vaccination with Ad26.COV2.S leads to persistence of protective efficacy (11). On the flip side, there are other evidences that a single dose of Ad26.COV2.S may not be sufficient; for instance a higher incidence of breakthrough infections with the Delta variant has been observed in comparison to mRNA vaccinated in some U.S. states (12). Additionally, a reduced ability of Ad26.COV2.S to induce antibody responses was reported in immunocompromised individuals (13). As a result, it has been proposed that individuals vaccinated with Ad26.COV2.S should receive a second dose, similarly to the two dose regimen recommended for mRNA-based vaccines (BNT162b2 and mRNA1273) and the adenoviral vector-based vaccine (ChAdOx1 nCov-19).

Vaccine-induced protective efficacy is associated with the ability to induce neutralizing anti-Spike antibodies and Spike-specific T cells (14). Data in animal models and in healthy individuals vaccinated with the other adenoviral vector-based vaccine, ChAdOx1 nCov19, followed by BNT162b2 have shown that a heterologous vaccine boost enhances both cellular and humoral immunity (15–18). Data recently reported showed an ability of both homologous and heterologous boost after Ad26.COV2.S to increase Spike-specific antibodies, however a parallel analysis for cellular immunity was not performed (19).

Therefore, to gain a more comprehensive information on the best boosting strategy in individuals vaccinated with a single dose of Ad26.COV2.S, we studied here, the Spike-specific T and B cell immunogenicity after a homologous or heterologous second vaccination dose. We compared the results with those obtained from individuals vaccinated with a single or double dose of BNT162b2. The quantity and breadth of Spike-specific antibodies and T cells were studied. Collectively, our data show the enhanced immunogenicity of the heterologous boosting strategy in individuals primed with Ad26.COV2.S.

## Methods and Materials

### Subject details

We recruited 115 study participants who received different vaccination regimes against COVID-19. Details are presented in Table 1.

**Table 1:**
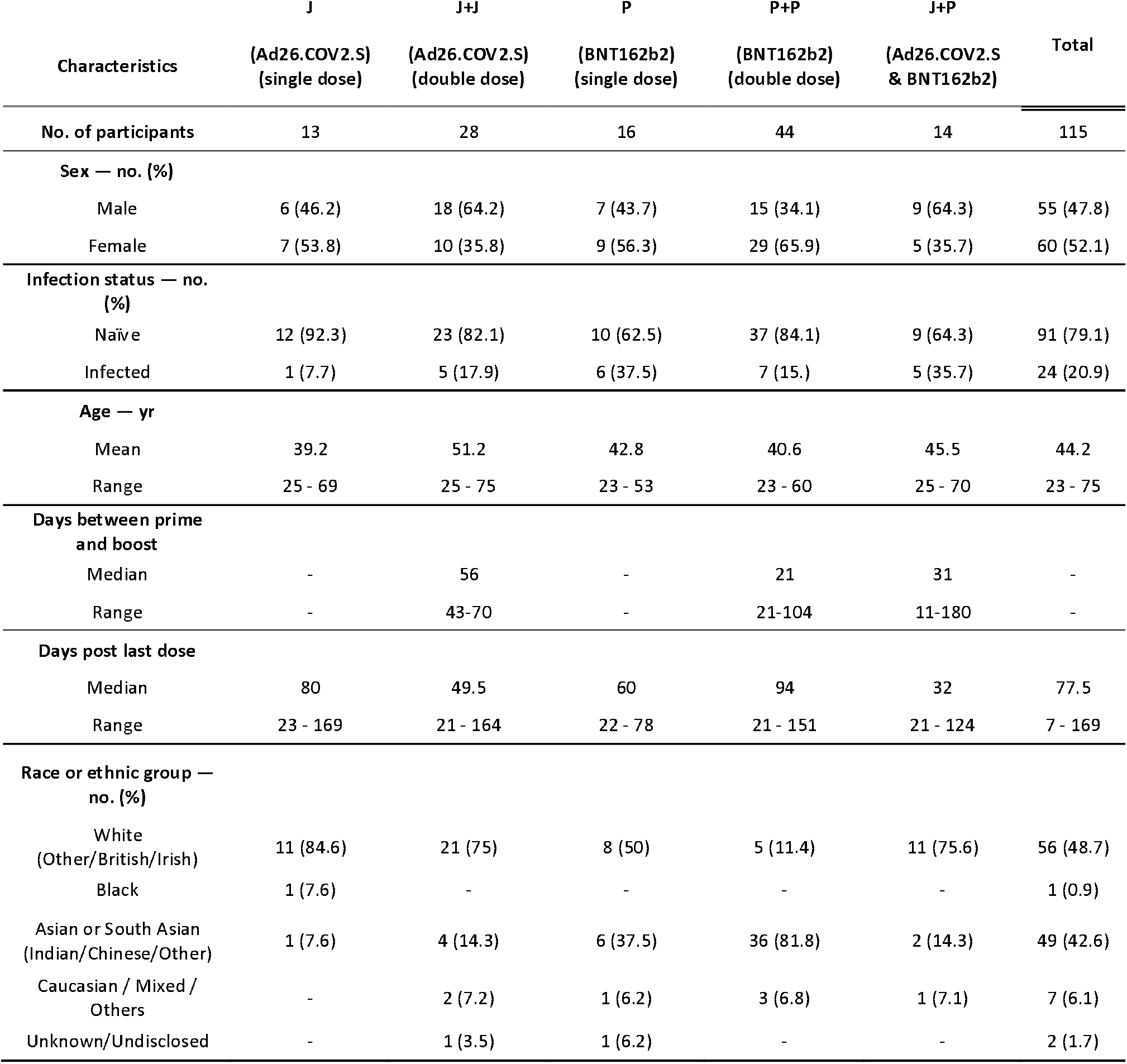
Demographics of vaccinated individuals

| Characteristics | J<br>(Ad26.COV2.S)<br>(single dose) | J+J<br>(Ad26.COV2.S)<br>(double dose) | P<br>(BNT162b2)<br>(single dose) | P+P<br>(BNT162b2)<br>(double dose) | J+P<br>(Ad26.COV2.S<br>& BNT162b2) | Total |
| --- | --- | --- | --- | --- | --- | --- |
| **No. of participants** | 13 | 28 | 16 | 44 | 14 | 115 |
| **Sex — no. (%)** |  |  |  |  |  |  |
| Male | 6 (46.2) | 18 (64.2) | 7 (43.7) | 15 (34.1) | 9 (64.3) | 55 (47.8) |
| Female | 7 (53.8) | 10 (35.8) | 9 (56.3) | 29 (65.9) | 5 (35.7) | 60 (52.1) |
| **Infection status — no. (%)** |  |  |  |  |  |  |
| Naïve | 12 (92.3) | 23 (82.1) | 10 (62.5) | 37 (84.1) | 9 (64.3) | 91 (79.1) |
| Infected | 1 (7.7) | 5 (17.9) | 6 (37.5) | 7 (15.) | 5 (35.7) | 24 (20.9) |
| **Age — yr** |  |  |  |  |  |  |
| Mean | 39.2 | 51.2 | 42.8 | 40.6 | 45.5 | 44.2 |
| Range | 25 - 69 | 25 - 75 | 23 - 53 | 23 - 60 | 25 - 70 | 23 - 75 |
| **Days between prime and boost** |  |  |  |  |  |  |
| Median | - | 56 | - | 21 | 31 | - |
| Range | - | 43-70 | - | 21-104 | 11-180 | - |
| **Days post last dose** |  |  |  |  |  |  |
| Median | 80 | 49.5 | 60 | 94 | 32 | 77.5 |
| Range | 23 - 169 | 21 - 164 | 22 - 78 | 21 - 151 | 21 - 124 | 7 - 169 |
| **Race or ethnic group — no. (%)** |  |  |  |  |  |  |
| White<br>(Other/British/Irish) | 11 (84.6) | 21 (75) | 8 (50) | 5 (11.4) | 11 (75.6) | 56 (48.7) |
| Black | 1 (7.6) | - | - | - | - | 1 (0.9) |
| Asian or South Asian<br>(Indian/Chinese/Other) | 1 (7.6) | 4 (14.3) | 6 (37.5) | 36 (81.8) | 2 (14.3) | 49 (42.6) |
| Caucasian / Mixed /<br>Others | - | 2 (7.2) | 1 (6.2) | 3 (6.8) | 1 (7.1) | 7 (6.1) |
| Unknown/Undisclosed | - | 1 (3.5) | 1 (6.2) | - | - | 2 (1.7) |

### Peripheral blood mononuclear cell isolation

Peripheral blood of all individuals was collected and peripheral blood mononuclear cells (PBMC) were isolated by Ficoll-Paque density gradient centrifugation.

### Peptides

15-mer peptides that are overlapping by 10 amino acids (AA) spanning the entire SARS-CoV-2 Spike protein, Nucleoprotein and Membrane protein were synthesized (Genscript) and pooled into 7, 2 and 1 pools of approximately 40 peptides in each pool, respectively (20, 21).

### SARS-CoV-2 Spike-specific T cell quantification

The frequency of SARS-CoV-2-specific T cells was quantified as described previously (22). Briefly, cryopreserved PBMCs were stimulated with peptide pools in an IFN-γ ELISpot assay. ELISpot plates (Millipore) were coated with human IFN-γ antibody overnight at 4°C. 400,000 PBMCs were seeded per well and stimulated for 18h with the distinct peptide pools at 2 μg/ml. The plates were then incubated with human biotinylated IFN-γ detection antibody, followed by Streptavidin-AP and developed using the KPL BCIP/NBT Phosphatase Substrate. To quantify positive peptide-specific responses, 2x mean spots of the unstimulated wells were subtracted from the peptide-stimulated wells, and the results expressed as spot forming cells (SFC)/10^6^ PBMC. Results were excluded if negative control wells had >30 SFC/10^6^ PBMC or if positive control wells (PMA/Ionomycin) were negative.

### Surrogate Virus Neutralization Test (sVNT)

The sVNT assay is a proxy measurement of antibodies inhibiting SARS-CoV-2 virus binding to the host cell receptor, human angiotensin-converting enzyme 2 (hACE2), and has been shown to correlate closely with antibody neutralization of SARS-CoV-2 (23). sVNT was measured using a commercial RBD-hACE2 binding inhibition assay called cPASS™ (GenScript) as per manufacturer guidelines. Briefly, serum was diluted 1:10 in the kit sample buffer, was mixed 1:1 with HRP-conjugated RBD and incubated for 30 mins at 37°C. RBD-antibody mixtures were then transferred and incubated for 15 mins at 37°C in enzyme-linked immunosorbent assay (ELISA) plates coated with recombinant hACE2 receptor. Following incubation, plates were washed with wash solution, incubated with TMB substrate for 12-15 mins and reaction stopped with stop solution. Absorbance was measured at OD_450nm_. Percent inhibition of RBD-hACE2 binding was computed using the following equation:

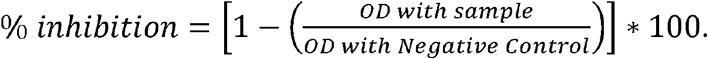

### SARS-CoV-2-specific Luminex Antibody assay

Antigen-specific IgG and IgA responses in serum samples were measured using a previously described bead-based immune-assay with some adjustments (22). Briefly, SARS-CoV-2 recombinant proteins Spike, S1 or S2 (AcroBiosystems) were covalently conjugated to Magpix Luminex beads. Antigen-conjugated beads were then blocked with 1% BSA (bovine serum albumin, before being probed with either diluted human serum or antibody standards for 1 hr at 37C. Beads were then washed and probed with either anti-human IgG-PE (Invitrogen) or anti-human IgA-Biotin (Southern Biotech) followed by Streptavidin-PE (Southern Biotech) for measuring human IgG and IgA, respectively. IgG and IgA binding to antigen were measured as Median Fluorescence Intensity (MFI) using a Magpix instrument (Luminex). MFI values of serum samples were converted to antibody quantity (i.e. g/ml) using anti-Spike IgG and IgA antibody standards (AcroBiosystems). Serum samples were first tested at dilutions 1:100 and 1:2000, if MFI values were above the range of the antibody standards, then serum samples were further diluted to 1:10,000 and tested.

### Detection of SARS-CoV2-specific Memory B cells

To detect SARS-CoV-2-specific memory B cells, biotinylated protein antigens were individually multimerized with streptavidin (SA) fluorophore conjugates, as described here (24). Briefly, full length Spike protein (RnD Systems) was multimerized with SA-Dylight 550 or SA-Dylight 650 (Thermo Fisher Scientific) in buffer containing 50/50 mixture of 2% FBS and Brilliant Buffer (BD Bioscience) for 1 hour at 4°C. Spike protein (RnD Systems) and SA-Dy550/SA-Dy650 were mixed at a 10:1 mass ratio (∼4:1 molar ratio) freshly before every staining. Cells were first stained for 10 minutes at RT with Live/Dead Fixable Blue Stain Reagent (Life Technologies). Subsequently cells were stained with 50µl of antigen probe cocktail containing 100ng of Spike per probe (i.e. 100ng of Spike-Biotin/SA-Dylight 550 and 100ng of Spike-Biotin/SA-Dylight 650) for 1 hour at 4°C. In parallel, SA-Dylight 550 and SA-Dylight 650 probes (100ng each) not conjugated to protein were used as decoy probes to gate out non-specific streptavidin-binding B cells. Next, cells were stained with an antibody cocktail (against CD3, CD10, CD19, CD21, CD27, CD38, CD40, CD69, CD71, CD95, IgD, IgG, IgM, see Table S1) for 30 mins at 4°C. Finally, cells were washed and fixed with 1% formaldehyde before acquisition on a LSR-Fortessa flow cytometer (BD). Analysis of flow cytometry data was performed using FlowJo software, version 10 (BD).

### Study approval

All donors provided written consent. The study was conducted in accordance with the Declaration of Helsinki and approved by the NUS Institutional Review Board (NUS-IRB-2021-292), the SingHealth Centralised Institutional Review Board (CIRB ref.: 2018/2387; 2018/3045; 2021/2014) and the Queen Mary University of London Review Board (REC Ref: 20/EE/0154).

### Statistical analyses

All statistical analyses were performed in Prism (GraphPad Software); details are provided in the figure legends.

## Results

### Cohorts of vaccinated individuals

We studied humoral and cellular immunity in a total of 115 individuals who received different vaccination schedules (Fig. 1A): single dose Ad26.COV2.S (J; n=13), homologous double dose Ad26.COV2.S (J+J; n=28), single dose BNT162b2 (P; n=16), homologous double dose BNT162b2 (P+P; n=44) and heterologous switch dose Ad26.COV2.S followed by BNT162b2 (J+P; n=14). Epidemiological characteristics of the studied population are summarized in Table 1. As controls, we studied 10 to 22 unvaccinated healthy individuals and 40 unvaccinated SARS-CoV-2 convalescents. In the homologous double dose Ad26.COV2.S cohort, the median time between the first and second dose was 56 days (43-71 days). In the heterologous switch dose Ad26.COV2.S and BNT162b2 cohort, the median number of days between first and second dose was 31 days, but with a wider range (11-180 days). The time between first and second dose of BNT162b2 was 21 days (21-104 days). Note also that the analysis of humoral and cellular immune parameters was performed at variable intervals after the second dose with a median number of days indicated in Table 1. Humoral responses were characterized by measuring IgG and IgA against the whole Spike, S1 and S2 domains, neutralising antibodies (using the surrogate virus neutralization test; sVNT) and by quantification of Spike-specific memory B cells (Fig. 1B). T cell response was analysed by quantification of IFN-γ secreting cells in reaction to peptides covering the whole Spike protein in an ELISpot assay. Seven pools of 15-mer peptides were used to detect Spike-specific T cells, Spike pools 1 to 4 were derived from the S1 chain, including the signal sequence. Spike pools 5 to 7 were derived from the S2 chain (Methods, Fig. 1C).

**Figure 1:**
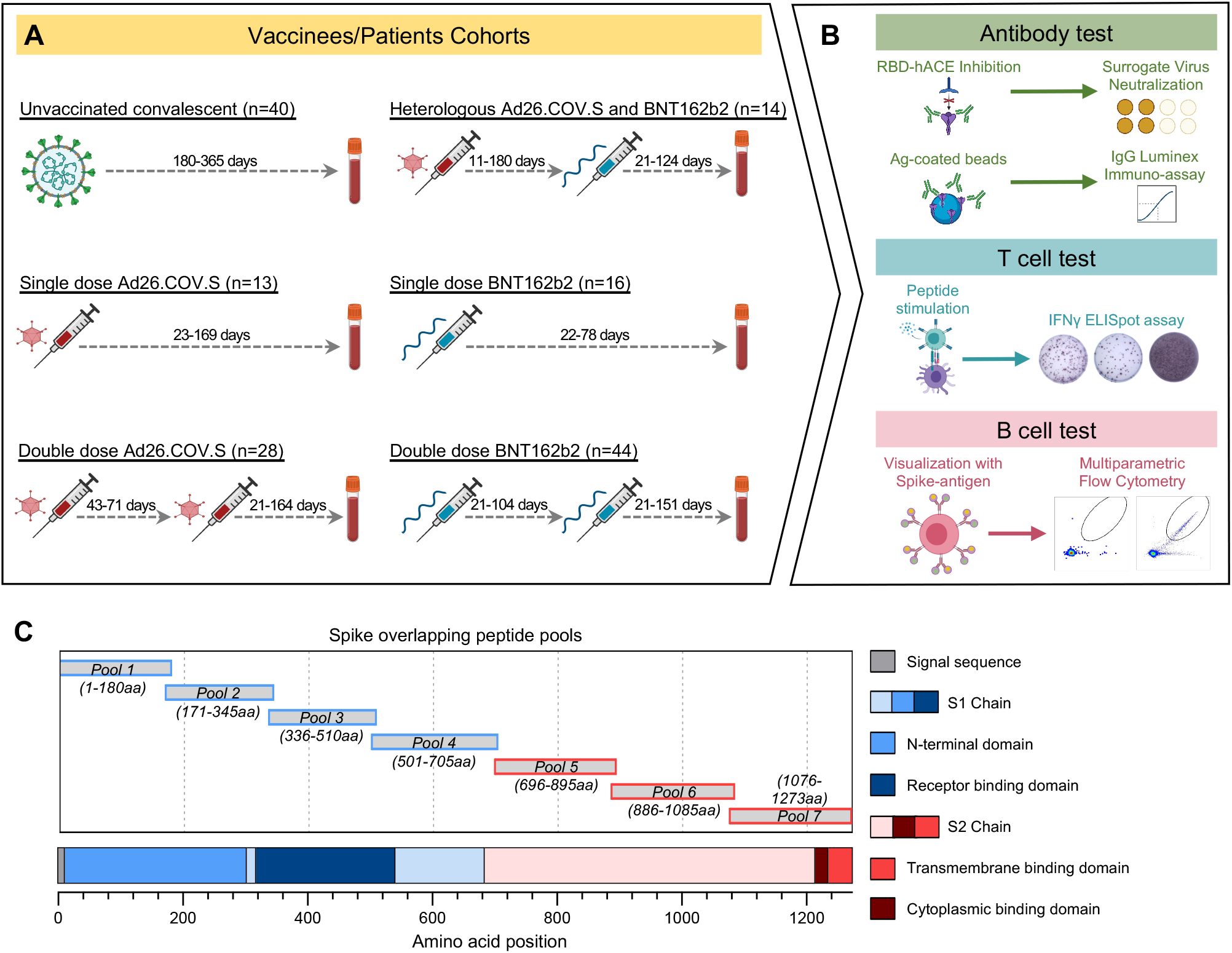
Vaccinated individuals and immune characterization. **(A)** Immune characterization was done in six cohorts: single dose Ad26.COV2.S (J), double dose Ad26.COV2.S (J+J), heterologous Ad26.COV2.S + BNT162b2 (J+P), single dose BNT162b2 (P) and double dose BNT162b2 (P+P). Unvaccinated cohort was used as a control (Unvac). Thenumber of individuals for each cohort, the interval between first and second dose of vaccine and the range of days for blood collection after last dose of vaccine are indicated. Sampling for immune analysis (humoral and cellular) was done only after the last vaccination dose both in homologous and heterologous vaccinated groups. **(B)** Schematic of humoral and cellular analysis. **(C)** Schematic representation of the location of the seven Spike-specific peptide pools containing 15-mer overlapping peptides spanning the entire Spike protein. Pools 1 to 4 contain peptides from the signal peptide and the S1 chain. Pools 5 to 7 encompass the S2 chain together with the transmembrane and cytoplasmic domains.

Furthermore, to ensure that we could differentiate individuals who had prior exposure to SARS-CoV-2 infection, presence of SARS-CoV-2 Membrane and Nucleoprotein-specific T cells were determined (21). 24 individuals tested positive for at least one of the three peptide pools used and they were classified as vaccination in SARS-CoV-2 convalescents in further analyses (Fig. S1).

### Quantification of humoral and cellular Spike-specific immune responses

We first quantified Spike-specific humoral and cellular immune responses in the five groups of vaccinated naïve individuals (Fig. 2A). Vaccine boost increased the overall profile of humoral immune responses in all individuals irrespective of their first vaccination (J or P). In individuals vaccinated with Ad26.COV2.S both homologous (J+J) and heterologous (J+P) vaccination increased the quantity of total Spike IgG and neutralising antibodies (sVNT). However, heterologous (J+P) vaccination induced a higher quantity of anti-Spike IgG and IgA antibodies than homologous (J+J) vaccination (Fig. 2A and S2A). The level of neutralising antibodies was also higher in heterologous versus homologous vaccinated individuals even though it did not reach statistical significance. Remarkably, all heterologous vaccinated individuals (9/9) had neutralising antibodies that achieved more than 80% of inhibition in the sVNT. In contrast, 7/21 of homologous vaccinated (J+J) had sVNT levels below 50%. Analysis of the frequency of circulating Spike-specific memory B cells was instead poorly indicative of the level of antibodies detected. Many individuals showed no or minimal increase of Spike-specific memory B cell frequency after booster, yet individuals with higher frequency were among the heterologous (J+P) vaccination cohort (Fig. 2A).

**Figure 2:**
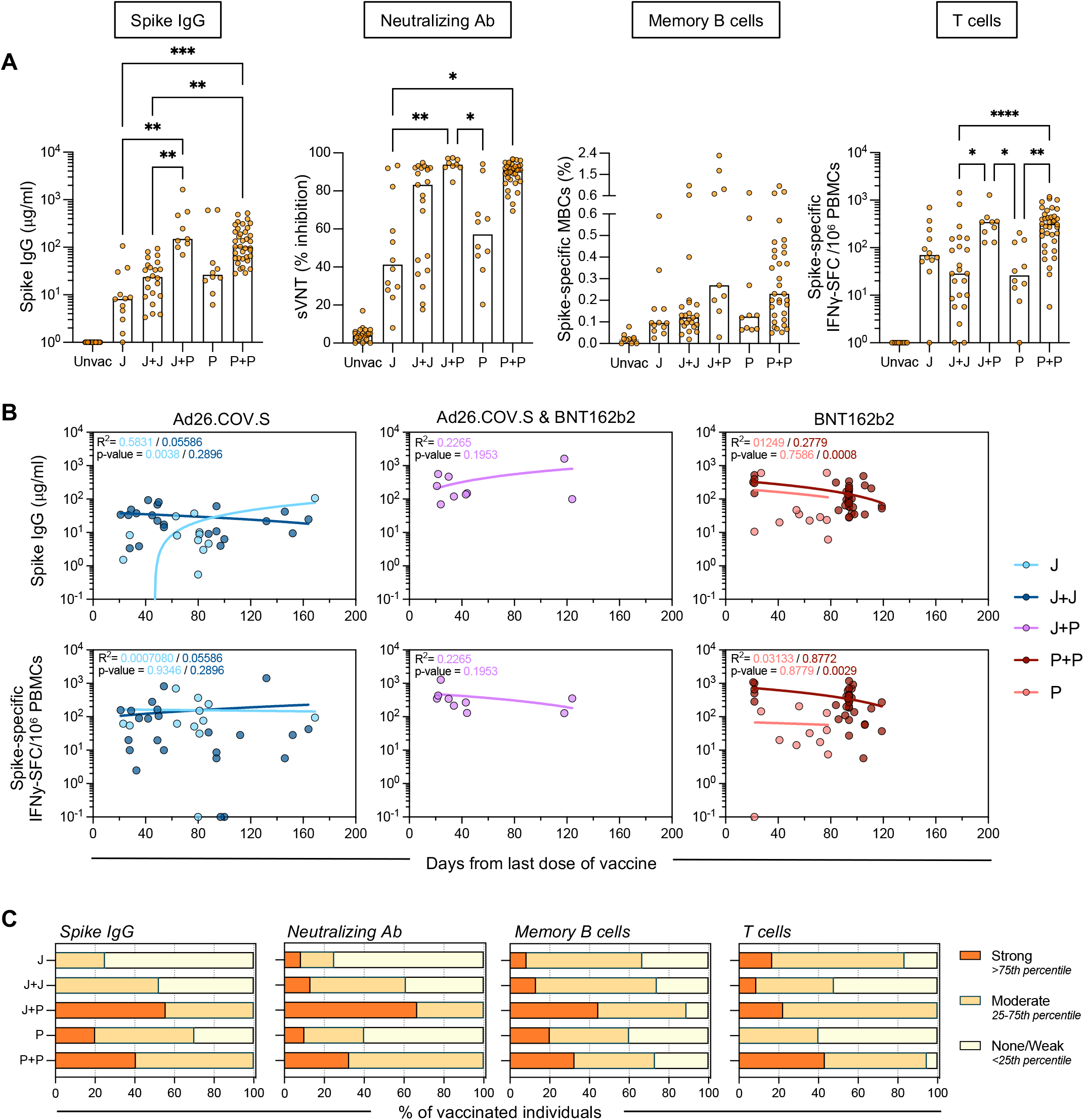
Quantification of humoral and cellular Spike-specific immune responses. **(A)** Spike IgG antibody, sVNT, Spike-specific MBCs and T cell responses were tested in five cohorts of naïve vaccinated individuals: J (n=12), J+J (n=23), J+P (n=9), P (n=10) and P+P (n=37). A naïve unvaccinated cohort was used as a control, Unvac (n=10-22). Bars denote the median value of each group. Each dot represents an individual. Significant differences in each group were analysed by one-way ANOVA and the adjusted p-value (adjusted for multiple comparison) are shown. No significance is not shown, * = P≤0.05; ** = P≤0.01; *** = P≤0.001; **** = P≤0.0001. **(B)** Linear regression analysis between neutralizing antibody activity (top panels) or Spike-specific T cell frequency (bottom panels) and time of testing after last vaccine dose (days). Goodness of fit and p-value ar4e shown in plots. **(C)** Bar graphs show the proportion of vaccinees with varying levels of Spike-IgG antibodies, neutralizing antibodies, Spike-specific B and T cell frequencies. The type of responders (Strong/moderate/none-weak) was expressed as a fraction of the number of vaccine recipients in each cohort. Type of responders were determined by percentile score calculated from all the vaccinees (n=87-91).

The analysis of Spike-specific T cells showed a higher frequency in heterologous (J+P) versus homologous (J+J) vaccine recipients (347.5 vs 152 SFC/10^6^ PBMC). This was similar to the level observed in double dose BNT161b2 vaccinated individuals. Notably, while Spike-specific T cells were clearly detected in 11/12 individuals vaccinated with a single dose Ad26.COV2.S, 10/23 individuals who received a homologous (J+J) booster displayed a weak level of Spike-specific T cells (<30 SFC/10^6^ PBMC) with a frequency similar to that observed in single dose BNT162b2 vaccinated individuals (Fig. 2A).

To ensure that differences in time of sampling after vaccination did not interfere with the observed trends, we investigated the effect of time after vaccination on immunogenicity by plotting sVNT and the frequency of Spike-specific T cells against the day of testing after the last dose of vaccine (Fig. 2B). Overall, as has been reported in previous studies, the Spike-specific T cell frequency and neutralizing antibodies in vaccinated individuals were not significantly reduced over time at least in the first 3-4 months post-vaccination (20, 25). Importantly, even though we tested the majority of individuals boosted with heterologous (J+P) vaccine within 30-40 days, Spike IgG quantity and T cell frequency remained high in the individuals tested at day 120 post second dose (Fig. 2B). Moreover, the individuals vaccinated with a single dose of Ad26.COV2.S or with homologous boost (J+J) displayed a pattern of Spike IgG and T cells that was not influenced by the time of testing (Fig. 2B). We also analysed Spike-specific T cell frequency in relation to the age and sex of the vaccine recipients (Fig. S3). Undetectable/low frequency of Spike-specific T cells was observed in the homologous vaccinated individuals who were above the age of 50. However, individuals of similar age (above 50 years old) present in the heterologous (J+P) vaccination cohort, all displayed high frequency of Spike-specific T cells (>100 SFC/10^6^ PBMC). Finally, the level of antibodies and T cell response to the different vaccine regimen were categorized into none/weak, moderate or strong responders based on their percentile ranking (Fig. 2C). The heterologous (J+P) vaccination cohort had the highest proportion of strong responders both in antibodies and T cell response. Instead, the homologous (J+J) group had the highest proportion of weak responders for T cells, lower than in single dose Ad26.COV2.S and equivalent to single dose BNT162b2 vaccinated individuals.

### Qualitative analysis of humoral and cellular Spike-specific immune responses

The ability to produce a polyclonal antibody response targeting different regions of the Spike protein is essential to maintain the protective efficacy of humoral immunity against SARS-CoV-2 variants (14, 26, 27). Similarly, T cell responses targeting multiple sites of Spike will reduce the chances of viral variants escaping T cell recognition (14, 28). Therefore, we analysed qualitative aspects of both humoral and cellular immunity induced by the different vaccination strategies.

First, we analysed the breadth of antibody responses. Antibodies (IgG and IgA) against the S1 (containing RBD) and the S2 regions of Spike were quantified. Homologous or single dose BNT162b2 vaccination elicited antibodies targeting both chains of Spike, while Ad26.COV2.S vaccination mounted an antibody response targeting preferentially the S1 chain (Fig. 3A, S2B). However, heterologous (J+P) vaccination appears to broaden the antibody repertoire against Spike since all the heterologous vaccinated individuals (9/9) had antibodies against both domains, while only 2/23 of homologous (J+J) vaccinated individuals displayed such antibody diversity (Fig. 3B, S1B).

**Figure 3:**
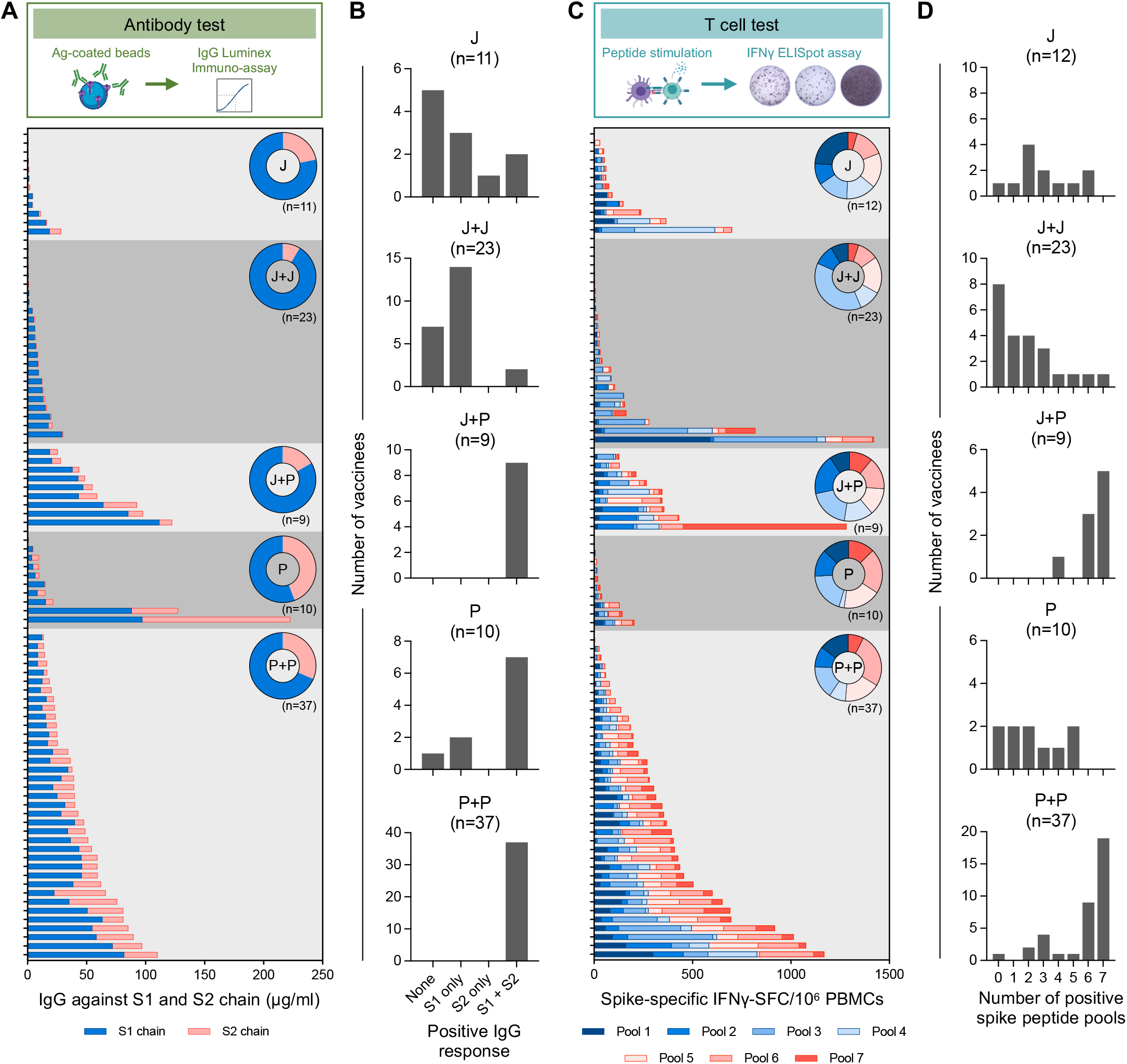
Qualitative profile of Spike-specific humoral and cellular immune responses. **(A)** Stacked bars represent IgG antibody titers against the S1 (blue) and S2 (pink) chains of SARS-CoV-2 Spike antigen. Each column represents an individual. Donut plots represent the mean of percentage of IgG antibodies against S1 or S2. **(B)** Frequency of with Spike-specific IgG antibodies recognizing none, S1, S2 or both S1 and S2 regions of Spike. IgG titers >1.35μg/ml were considered positive. **(C)** Stacked bars represent frequency of IFN-γ-spot forming cells (SFC) reactive to the individual Spike peptide pools (1 to 7) in each vaccinee. Donut chart represent the percentage mean of IFN-γ-SFC reactive to the individual Spike peptide pools. (**D**) Bar graphs show the frequency of vaccinees with varying breadth of Spike-specific T cell response, determined by the number of positive Spike peptide pools. Responses >7.5 SFC/10^6^ PBMCs were considered positive.

Also the analysis of the breadth of the Spike-specific T cells confirmed the ability of heterologous vaccination to broaden the immune response against Spike. Figure 3C shows the frequencies of Spike-specific T cells reactive to the distinct peptide pools covering the different regions of Spike (Fig. 1C). While heterologous (J+P) vaccination expanded a population of T cells able to recognize both S1 and S2 regions of Spike (a pattern similar was observed in the homologous BNT162b2 vaccination group), homologous (J+J) and single dose (J) vaccination induced T cells primary targeting the S1 chain.

The number of Spike-peptide pools recognised by T cells was also different (Fig. 3D). Individuals on the J+P vaccine regimen had T cells recognizing at least four Spike-specific peptide pools. Furthermore, 8/9 had highly multi-specific T cells recognizing six or seven different peptide pools. In contrast, a homologous Ad26.COV2.S booster did not expand the ability of Spike-specific T cells to recognize different regions (Fig. 3D). Only 4/23 individuals on homologous J+J had T cells recognizing at four or more distinct regions of Spike. A similar pattern was observed in single dose Ad26.COV2.S vaccinated individuals. Of concern, 8/23 homologous (J+J) vaccinated individuals did not develop a T cell response against any region of Spike (<7.5 SFC/10^6^ PBMC). In contrast, this was observed only 1/12 single dose Ad26.COV2.S vaccine recipients.

### Class switching of Spike-specific memory B cells

High affinity antibodies are produced by memory B cells (MBCs) upon re-encounter with viral antigen (29, 30). Particularly, class-switched IgG+ MBCs are responsible for durable humoral responses (31, 32). Having observed that a subset of homologous J+J vaccine recipients had none/weak antibody and T cell responses (Fig. 2C), this prompted us to analyse the profile of Spike-specific B cell maturation.

The proportion of class-switched IgG+ Spike-specific MBCs is shown in Figure 4. In all the cohorts we observed a vast heterogeneity in the proportion of class-switched MBCs among the individuals. Both homologous (J+J; P+P) and heterologous (J+P) booster vaccination increased the proportion of class-switched Spike-specific MBCs in comparison to a single dose of Ad26.COV2.S (J) or BNT162b2 (P) (Fig. 4B). However, there was no significant difference observed between homologous and heterologous boosting (Fig. 4C).

**Figure 4:**
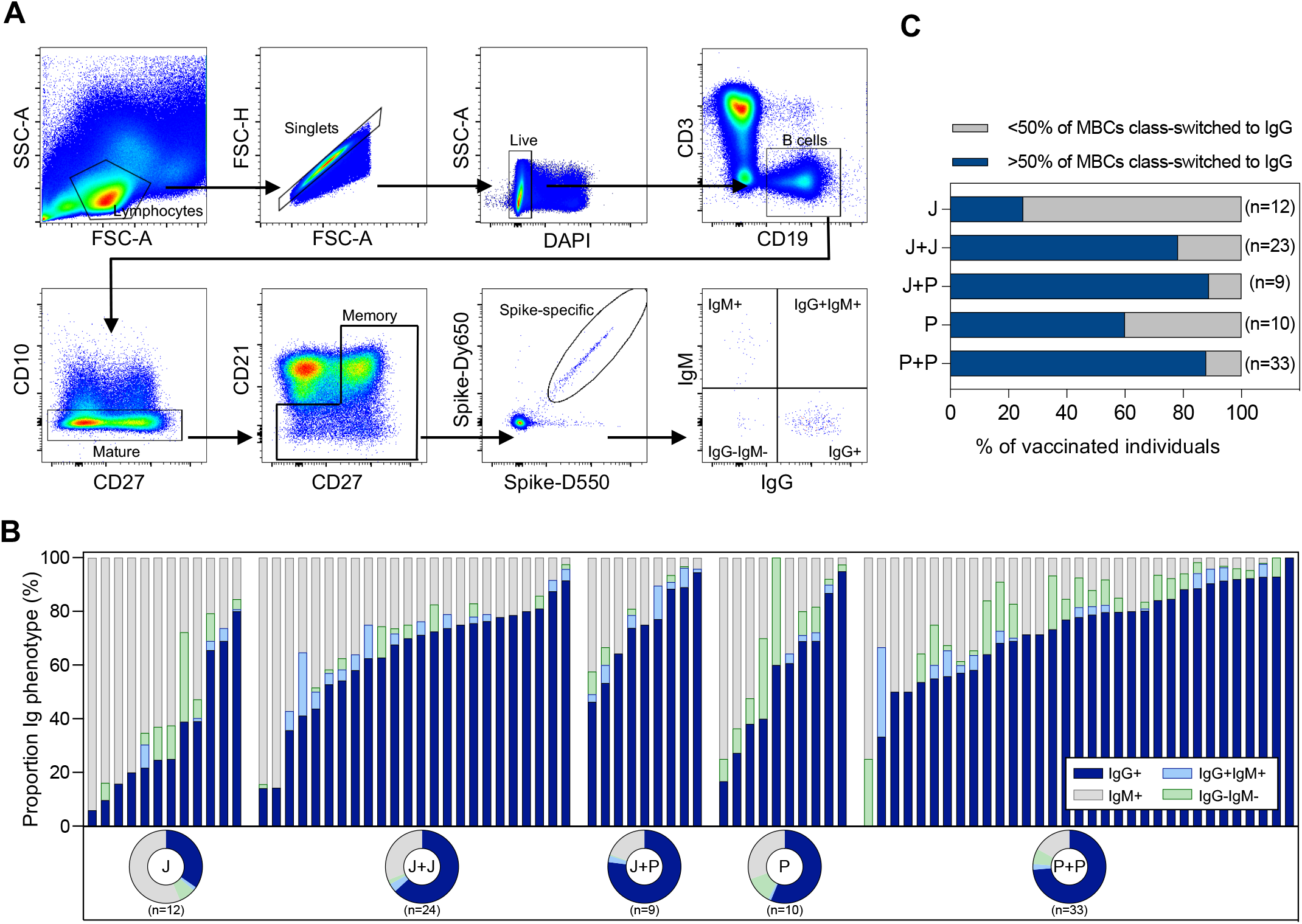
Phenotypic analysis of immunoglobulin isotypes of Spike-specific memory B cells. **(A)** Representative gating strategy of Spike-specific memory B cells (MBCs) expressing different immunoglobulin (Ig) types. **(B)** Stacked bars represent the frequency of indicated Ig isotypes expressed on Spike-specific MBCs. Corresponding donut plots represent the mean proportion of the four Ig categories (IgG+, IgG-IgM-, IgG+IgM+ and IgM+). **(C)** Frequency of individuals with >50% of MBCs class-switched to IgG (blue).

### Characterization of Spike-specific humoral and cellular immune responses in vaccinated SARS-CoV-2 convalescents

Presence of SARS-CoV-2 Membrane and Nucleoprotein-specific T cells allowed us to identify a total of 23 individuals who were vaccinated with different regimens but were likely infected by SARS-CoV-2 before or during the vaccination regimens (21). The limited number of these individuals categorized within the different vaccination strategies did not allow us to perform statistically significant comparisons. Note, that in the cohort of single dose Ad26.COV2.S (J) there was only one convalescent, hence any measurement of “possible boosting effect” by the second dose cannot be performed. Nevertheless, we compared the quantity of antibodies (total anti-Spike IgG, neutralizing antibodies sVNT) and Spike-specific T cells induced by the different vaccination regime in naïve versus convalescent individuals (Fig. 5A). We observed that all convalescent individuals elicited a stronger humoral and cellular immunity than naïve individuals irrespective of the vaccination regime (Fig. 5A), in line with recent data (33–35).

**Figure 5:**
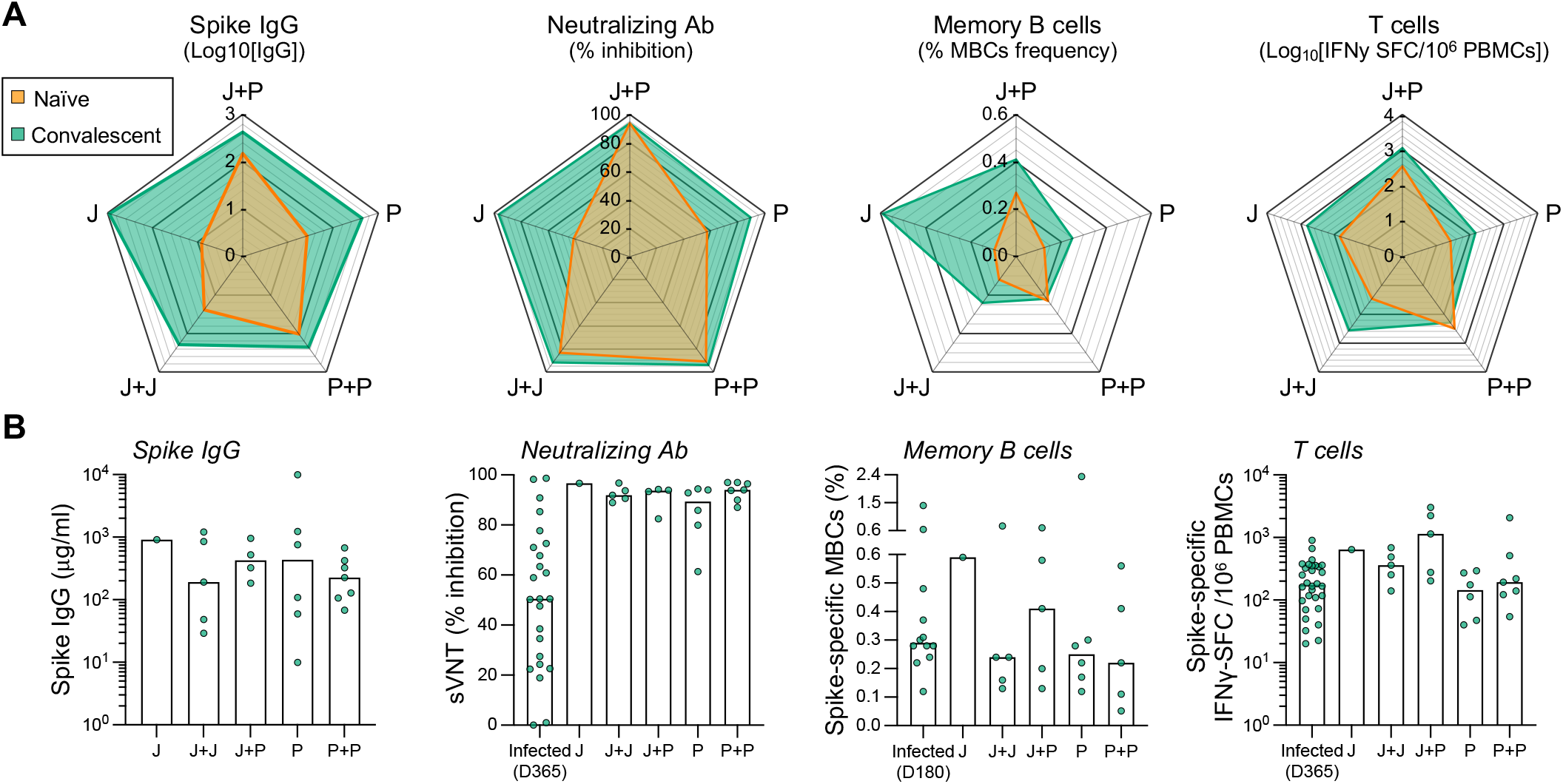
Spike-specific humoral and cellular immune response in convalescent vaccinated individuals. Spike IgG antibody, sVNT, Spike-specific MBCs and T cell responses were tested in five cohorts of naïve vaccinated individuals: J (n=12), J+J (n=23), J+P (n=9), P (n=10) and P+P (n=37). Same analysis was carried out in SARS-CoV-2 convalescents vaccinated with: J (n=1), J+J (n=5), P (n=6), P+P (n=7) and J+P (n=5). A convalescent unvaccinated cohort was used as a control: Infected (n=10-30). **(A)** The four radar plots show respectively median levels of Spike IgG antibody titers, neutralizing antibodies, frequency of Spike-specific memory B cells and T cells in convalescent (green; n=24) and naïve (orange; n=87-91) vaccinated individuals. Vaccination strategies (J, J+J, J+P, P and P+P) were indicated in the vertex of each pentagon. **(B)** Quantities of Spike IgG antibody, neutralizing antibodies, spike-specific memory B cells and T cells from convalescent vaccinated individuals. Bars denote the median value of each group. Each dot represents an individual. Significant differences in each group were analysed by one-way ANOVA and the adjusted p-values (adjusted for multiple comparison) are shown. No significance is not shown; * = P≤0.05; ** = P≤0.01; *** = P≤0.001; **** = P≤0.0001.

In addition, the different vaccination strategies in convalescent individuals boosted neutralizing antibodies above 90% inhibition, in comparison to the 50% inhibition present in non-vaccinated convalescents 1 year post infection. In contrast, their efficacy in boosting cellular immunity was minimal. The frequency of Spike-specific T cells was marginally increased (J+J) or even lower (P and P+P) than their level detected in unvaccinated convalescent individuals (Fig. 5B). However, high frequency of Spike-specific T cells was detected in heterologous vaccinated individuals. 3/5 individuals in this group displayed a quantity of Spike-specific T cells exceeding 1000 SFC/10^6^ PBMC.

## Discussion

Our study provides information on how to boost Spike-specific humoral and cellular immunity in individuals vaccinated with a single dose of Ad26.COV2.S.

Heterologous booster vaccination with a dose of BNT162b2 resulted in elevated titers of anti-Spike IgG and neutralizing antibodies and high frequency of Spike-specific T cell in all the individuals tested.

In addition, heterologous vaccination expanded the breadth of both humoral and T cell immunity. The results of Spike-specific T cells were particularly robust since we observed that 8/9 of the heterologous vaccinated individuals possessed T cells widely scattered among the whole length of the Spike protein. The immunological correlates of protection induced by the vaccines are still only hypothesized (36). Thus, we cannot conclude that the enhanced immunogenicity of heterologous vaccination will translate in a superior protective efficacy. However, it is likely that future prospective trials of vaccine efficacy will confirm that quantity and quality of humoral and cellular immunity might directly translate into protective efficacy against SARS-CoV-2 infection and COVID-19 disease.

Our observation that a heterologous vaccination strategy in Ad26.COV2.S vaccinated individuals is more immunogenic than a homologues boost was also confirmed in convalescents and is in line with the results obtained in individuals vaccinated with ChadOx1 nCov-19, another adenoviral-based vaccine. Heterologous vaccination after a single dose of ChadOx1 nCov-19 was recommended in several countries due to the problem of intermittent supply and of rare thrombotic events associated with ChadOx1 nCov-19 (37). Analysis of immunogenicity in individuals boosted with mRNA-based vaccines after a single dose of ChadOx1 nCov-19 revealed an enhanced quantitative profile of antibody and T cell response compared to homologous booster vaccination (15, 16). The superior immunogenicity of heterologous vaccination with a combination of vaccines utilizing different expression vectors (38) has also been seen in other vaccination strategies against different viruses (EBOLA, HIV, HBV (39–41)) and other pathogens (malaria, TB (42, 43)). This further suggests that heterologous prime-boost vaccination strategies involving different types of vaccine should be considered. For instance, using mRNA-based followed by an adenoviral-vector based vaccine might be able to delay the reduction of anti-Spike humoral immunity observed after homologous mRNA vaccination (5).

Importantly, although we observed that homologous vaccination with Ad26.COV2.S enhances the quantity of antibody production, only heterologous boosting expanded the ability of antibodies to recognise the S2 region of Spike. In addition, a second dose of Ad26.COV2.S did not enhance the quantity and breadth of Spike-specific T cells, and appeared detrimental in some individuals. More than one third (8/23) of individuals boosted with homologous Ad26.COV2.S vaccine did not display detectable Spike-specific T cell responses. This proportion of individuals with a very week T cell response was higher than in individuals who received a single dose of Ad26.COV2.S (1/12).

The inability of a homologous second dose Ad26.COV2.S to boost cellular immunity was also observed in non-human primates (44) as well as in individuals receiving homologous ChadOx1 nCov-19 vaccination (16, 17). However, the low level of Spike-specific T cells detected here in the individuals with Ad26.COV2.S homologous boost was unexpected and call for a careful evaluation of such strategy in individuals who already received a single dose of Ad26.COV2.S. In animals vaccinated with two doses of Ad26.COV2.S, the Spike-specific T cell response was found to be more stable (44). A parallel analysis of both humoral and cellular immune parameters in larger group of Ad26.COV2.S vaccinated individuals that are followed longitudinally is needed.

The discrepancy between levels of Spike-specific humoral and cellular immune responses has been also frequently observed in SARS-CoV-2 convalescents and vaccine recipients (reviewed in (45)). If we exclude the early phases of convalescence and vaccination, the level of Spike-specific antibodies and T cells appears independently regulated in numerous studies (20, 25, 46). As such, the data gathered here further support the concept that immunogenicity of vaccines should be comprehensively evaluated in its cellular and humoral branch. Humoral analysis alone cannot be used to evaluate the induction of virus-specific T cells.

There are some important limitations in our study. In addition to the small sample size, the cross-sectional nature of the study did not allow to precisely evaluate the modification of Spike-specific T cell frequency induced by the second dose at individual level. Furthermore, the analysis of both humoral and cellular immunity was performed at different time points after vaccination. Even though we showed that the level of Spike-specific T cells was minimally reduced within the first 6 months after vaccination and did not appear to influence the different pattern of Spike-specific T cells, an analysis at identical time points after boosting is indicated to demonstrate the enhanced immunogenicity of heterologous vaccination and its durability over time. Finally, the limited quantity of PBMCs collected only allowed us to perform a simple ELISpot analysis of the T cell response, a method that cannot discriminate whether the Spike-specific T cells induced by different vaccination regimens are CD4 or CD8 T cells. Such information would be needed to better characterize the possible further qualitative differences in T cells responses induced by the different vaccination regimens.

In conclusion, while the Ad26.COV2.S vaccine has been initially proposed as a single dose vaccine, the progressive reduction of its protective efficacy against SARS-CoV-2 infection over time have warranted a better definition of the best boosting strategy (12). Here we provide data that demonstrate the enhanced immunogenicity of heterologous versus homologous boost vaccination in Ad26.COV2.S vaccine recipients. Despite representing a minority within the vaccinated individuals worldwide, the Ad26.COV2.S vaccine recipients deserve information that can guide their future vaccination choice.

## Data Availability

All data produced in the present study are available upon reasonable request to the authors.

## Acknowledgments

We would like to thank all clinical and nursing staff who provided care to all vaccinated individuals and aided in co-ordinating collection of the samples. We are also grateful to all clinical trial coordinators and staff at Barts Vaccine centre for assistance in recruitment organisation of vaccine recipients.

---

**Supplementary Table 1:** List of antibodies used for Spike-specific B cell phenotyping

| Antibody | Catalogue Number | Concentration |
| --- | --- | --- |
| Brilliant Violet 605™ anti-human CD3 Antibody | 317322 | 2:50 |
| PE-CF594 Mouse Anti-Human CD10 | 562396 | 2:50 |
| BV510 Mouse Anti-Human CD19 | 562947 | 2:50 |
| BV421 Mouse Anti-Human CD21 | 562966 | 1:50 |
| BV650 Mouse Anti-Human CD27 | 563228 | 3:50 |
| PE/Cyanine7 anti-human CD38 Antibody | 356608 | 2:50 |
| PerCP/Cyanine5.5 anti-human CD40 Antibody | 334316 | 2:50 |
| FITC anti-human CD71 Antibody | 334104 | 3:50 |
| APC-Cy™ 7 Mouse Anti-Human CD69 | 557756 | 1:50 |
| BUV737 Mouse Anti-Human CD95 | 564710 | 1:50 |
| Alexa Fluor® 700 anti-human IgD Antibody | 348230 | 2:50 |
| BV786 Mouse Anti-Human IgG | 564230 | 2:50 |
| Brilliant Violet 711™ anti-human IgM Antibody | 314540 | 2:50 |
| LIVE/DEAD™ Fixable Blue Dead | L23105 | 2:1000 |

**Supplementary Figure 1:**
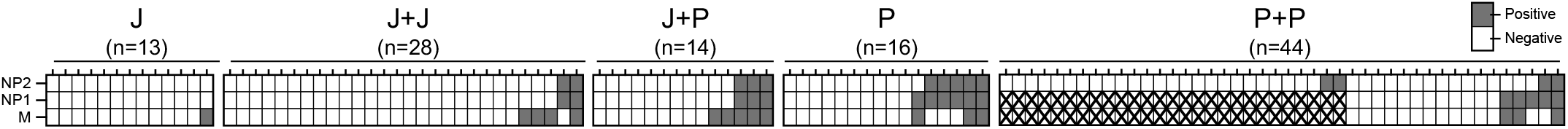
Identification of convalescent vaccinated individuals. Heatmap shows the SARS-CoV-2 non-Spike-specific T cell responses quantified in vaccinated individuals using IFN-γ ELISpot. Individuals with at least one positive response (>20 SFC SFC/10^6^ PBMCs) are categorized as potentially SARS-CoV-2 convalescent vaccinees. While individuals with no positive response are categorized as naïve vaccinees. “X” denotes structural peptide pools that were untested.

**Supplementary Figure 2:**
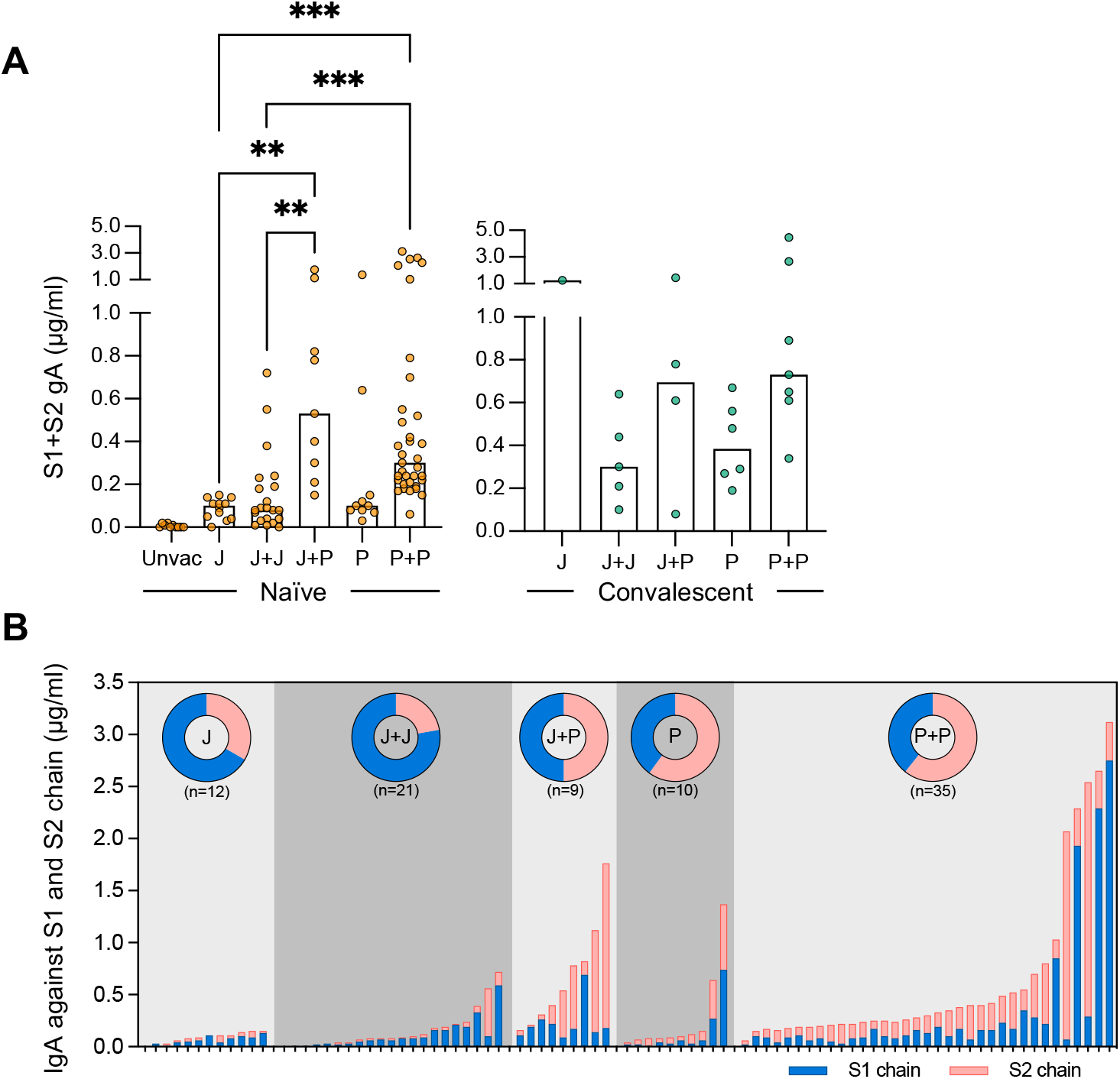
Quantitative and Qualitative profile of Spike-specific IgA antibodies. **(A)** Spike-specific IgA antibody responses were tested in five cohorts of naïve vaccinated individuals: J (n=12), J+J (n=23), J+P (n=9), P (n=10) and P+P (n=37). Same analysis was carried out in convalescent vaccinated individuals: J (n=1), J+J (n=4), J+P (n=5), P (n=6) and P+P (n=7). A naïve unvaccinated cohort was used as a control, Unvac (n=10). Bars denote the median value of each group. Each dot represents an individual. Significant differences in each group were analysed by one-way ANOVA and the adjusted p-values (adjusted for multiple comparison) are shown. No significance is not shown, * = P≤0.05; ** = P≤0.01; *** = P≤0.001. **(B)** Stacked bars represent IgA antibody titers against the S1 (blue) and S2 (pink) chains of SARS-CoV-2 Spike antigen. Each column represents an individual. Donut plots represent the mean of percentage of IgA antibodies against S1 or S2.

**Supplementary Figure 3:**
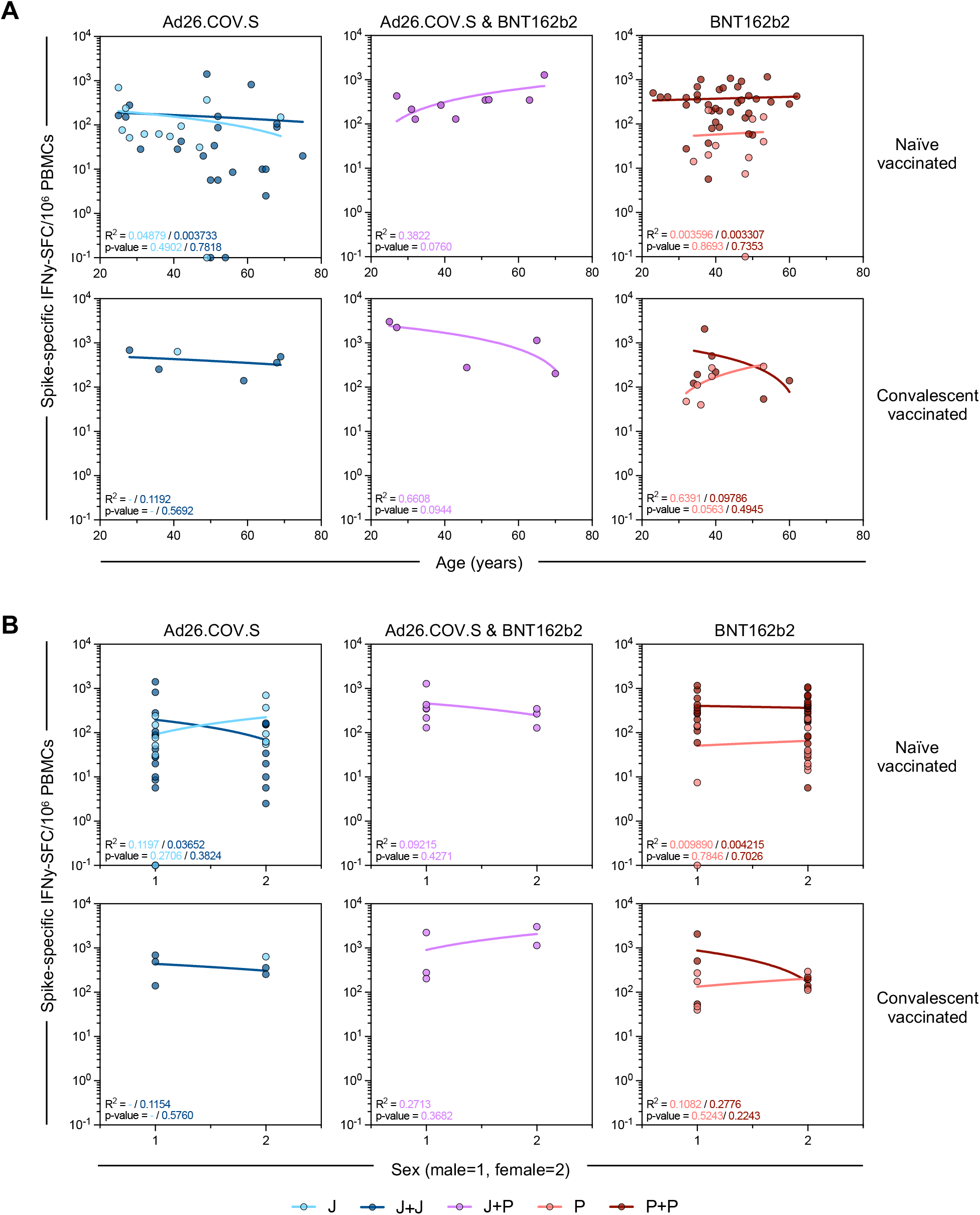
Correlation of Spike-specific T cell frequency with age or sex. Linear regression analysis between Spike-specific T cell frequency and **(A)** age or **(B)** sex of vaccine recipients who are naïve (top panel) or convalescents (bottom panel). Goodness of fit and p-value are shown in plots.

